# The effect of prenatal multiple micronutrient supplementation on birth weight in Ethiopia: protocol for a pragmatic cluster-randomised trial

**DOI:** 10.1101/2024.10.07.24315025

**Authors:** Tanya Marchant, Senait Alemayehu, Elias Asfaw, Alemneh Kabeta Daba, Atkure Defar, Charles Opondo, Lars Åke Persson, Joanna Schellenberg, Girum Taye, Anene Tesfa, Bedasa Tessema, Kalkidan Zenebe, Masresha Tessema

## Abstract

**Introduction:** This programme effectiveness study responds to the need for evidence of the effect on birth weight of switching from iron-folic acid supplementation to multiple micronutrient supplementation as part of routine antenatal care in Ethiopia. A 2019 meta-analysis reported a mean increase of 35 grammes in birth weight amongst newborns of women who took multiple micronutrient supplements in pregnancy compared to those who took iron-folic acid. Responding to that evidence, the government of Ethiopia decided to implement a 21 district pilot of the use of multiple micronutrient supplementation in routine antenatal care and requested an evaluation of implementation outcomes, including on birth weight.

**Methods and analysis:** A pragmatic, facility-based, randomised trial is being conducted in 42 districts over 5 regions of Ethiopia between January 2023 and December 2024. Districts have been randomised to one of two arms, either to retain iron-folic acid supplementation as part of routine antenatal care or switch to multiple micronutrient supplementation. During the study period the birth weights of all eligible babies born in enrolled health facilities in these 42 districts are continuously recorded alongside data on maternal receipt and use of either multiple micronutrient or iron-folic acid supplementation. We hypothesise that newborns of women resident in the 21 multiple micronutrient supplementation districts will have higher mean birthweight than newborns of women resident in the 21 iron-folic acid supplementation districts. Facility surveys involving pregnant women and health care workers at baseline, midline, and endline contribute to a process evaluation and cost and cost-effectiveness evaluation.

**Discussion:** Results from this pragmatic trial will be used by the government of Ethiopia in assessing success of the MMS pilot and for decision making about subsequent scale-up. The findings will also inform global decision making, particularly in settings where a transition from iron-folic acid to multiple micronutrient supplementation is being contemplated at scale.

**Trial registration number:** ClinicalTrials.gov Identifier: NCT05708183; protocol version 2

**Strengths and limitations of this study:**

- This pragmatic cluster-randomised trial evaluates implementation outcomes, processes and costs of a large-scale policy change to prenatal care piloted in Ethiopia, generating evidence that is actionable for country decision making and relevant to other settings.
- Continuous data collection, capturing information on all births in enrolled health facilities, makes use of and strengthens routine health management information systems.
- The design permits a dose-response analysis considering the changes in birth weight for different levels of coverage, and permits adjustment for seasonality.
- Interpretation of trial outcome findings must be made in the context of coverage of and adherence to prenatal MMS and IFA in the pilot areas.

## Introduction

Interventions to address maternal nutrition and low birth weight (LBW) include the optimal use of antenatal care (ANC) where micronutrient supplementation is provided as part of a package of essential pregnancy interventions.^1^ Traditionally, the provision of micronutrient supplementation at ANC has centred on iron-folate supplements (IFA) but there are advocates for a transition from IFA to multiple-micronutrient supplements (MMS).^2^ Numerous trials have investigated the extent to which use of MMS during pregnancy may improve maternal, foetal and infant health outcomes. In 2019, a meta-analysis of data from 19 MMS trials conducted in low- and middle-income countries concluded that, relative to IFA, prenatal use of MMS reduced the number of LBW newborns by 12% (pooled risk ratio 0.88, 95% confidence interval 0.85 to 0.91). Further, MMS was reported to lead to a 5% reduction in preterm births (pooled risk ratio 0.95, 95% confidence interval 0.90 to 1.01), and a 8% reduction in babies considered small-for-gestational age (pooled risk ratio 0.92, 95% confidence interval 0.88 to 0.97).^3^ In response to this meta-analysis, WHO updated its guideline on MMS to “recommended use in the context of rigorous research”.^1^

The government of Ethiopia is committed to improving health care so that more infants and children survive and thrive. One factor that contributes to poor child health in the country is the underlying poor nutrition of women and girls who frequently have multiple-micronutrient deficiencies.^4^ Persistent nutritional problems have life-cycle consequences with short, medium and long-term effects, including through the birthweight of newborns; 17% of Ethiopian newborns are estimated to be born low birth weight (<2500g).^5^

The Ministry of Health (MoH) of Ethiopia now wants to understand the effect on birth weight of a policy change from provision of IFA to provision of MMS as part of routine ANC. For this purpose, in 2023 MoH removed IFA supplements from the routine ANC package in 21 districts across five regions of Ethiopia so as to pilot the provision of MMS instead. The current pragmatic cluster-randomised trial aims to generate evidence on the effect on birth weight of this pilot policy change, in addition to providing evidence on implementation processes and costs.

## Methods and analysis

### Ethiopian context

Ethiopian health policy states that women should have eight antenatal care contacts during pregnancy with the first contact taking place in the first trimester of pregnancy, and that they should consume iron-folate supplements throughout pregnancy and for 180 days afterwards, and that childbirth should be attended by a skilled provider in a health facility.^6^ Table 1 shows a summary of the uptake of these recommendations between 2011 and 2019.^7^ Although considerable progress was made during the period, by 2019 most women did not attend clinic early in pregnancy or have the recommended number of antenatal visits, did not consume iron tablets for the recommended duration, and did not deliver in a health facility.^7^ This context of relatively low use of recommended care underlines the importance of understanding programme feasibility, the acceptability of interventions, and adherence to recommended schedules.

**Table 1.**
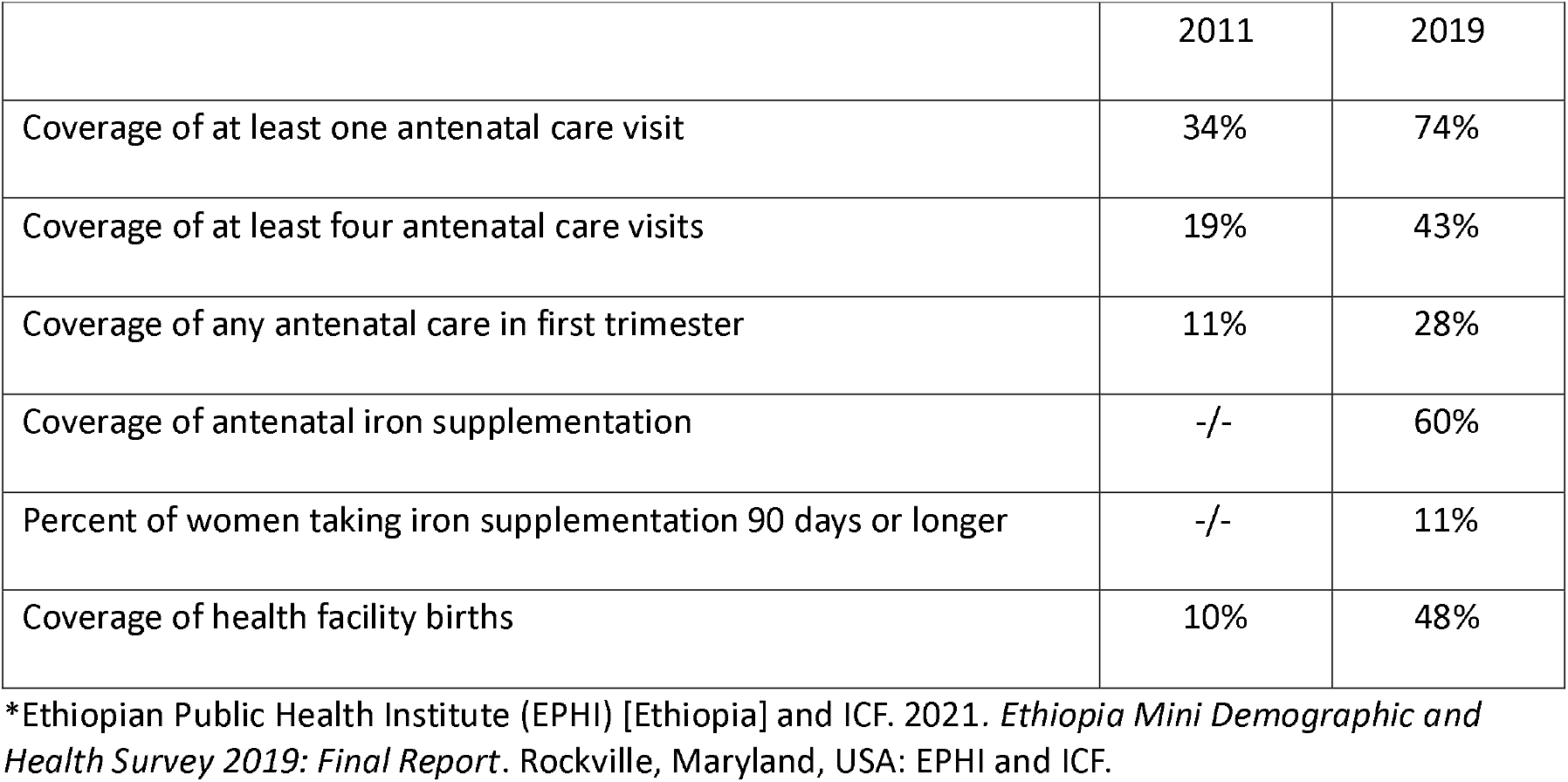
Population level coverage of antenatal care and health facility delivery in 2011 and 2019, Ethiopia Demographic and Health Surveys*.

|  | 2011 | 2019 |
| --- | --- | --- |
| Coverage of at least one antenatal care visit | 34% | 74% |
| Coverage of at least four antenatal care visits | 19% | 43% |
| Coverage of any antenatal care in first trimester | 11% | 28% |
| Coverage of antenatal iron supplementation | -/- | 60% |
| Percent of women taking iron supplementation 90 days or longer | -/- | 11% |
| Coverage of health facility births | 10% | 48% |

\*Ethiopian Public Health Institute (EPHI) [Ethiopia] and ICF. 2021. *Ethiopia Mini Demographic and Health Survey 2019: Final Report*. Rockville, Maryland, USA: EPHI and ICF.

### Study aim and objectives

The overall aim of the MMS evaluation is to investigate the programme effectiveness on birth weight of providing MMS as part of routine antenatal care, relative to providing IFA. The primary objective is to estimate the effect of MMS implementation on the mean birth weight of babies born in government health facilities to women living in areas where MMS is implemented, relative to the mean birth weight of babies born in government health facilities to women living in areas where standard antenatal IFA supplementation is implemented. This objective is addressed through *intention-to-treat* analysis.

The secondary objective is to estimate the effect of MMS on mean birth weight of newborns of mothers who receive and use either MMS or IFA. That is, a *per-protocol* analysis restricting to women who self-report having (i) received and (ii) consumed either IFA or MMS, with exploratory analysis around the number of tablets reported to be consumed. Additional secondary objectives are to investigate the processes of MMS implementation, how providers and pregnant women respond to MMS, and to estimate the cost and cost-effectiveness of MMS relative to IFA.

### Intervention details

With the support of UNICEF, the 21 district pilot policy shift in Ethiopia aims to substitute IFA for MMS as part of routine ANC. For MMS, the UNIMMAP (United Nations International Multiple Micronutrient Antenatal Preparation) formulation is used that contains 15 essential vitamins and minerals including: *Retinol (Vitamin A-acetate) 800 μg; Vitamin E (as vitamin E-acetate) 10 mg; Vitamin D (as Cholecalciferol) 200 IU; Vitamin B1 (Thiamine mononitrate) 1*.*4 mg; Vitamin B2 (As Riboflavin) 1*.*4 mg; Vitamin B3 (As Nicotinamide) 18 mg; Vitamin B6 (as Pyridoxine 1*.*9 mg; Vitamin B12 (as Cyanocobalamin) 2*.*6 mg; Folic Acid 400 μg; Vitamin C (As Ascorbic Acid) 70 mg; Iron (As ferrous sulphate) 30 mg; Zinc (As zinc sulphate) 15 mg; Copper (as Copper Sulphate) 2 mg; Selenium (as Sodium selenite) 65 μg; Iodine (as Potassium Iodate) 150 μg*.^8^

During 2023-2024, the goals of this pilot are to (1) create an enabling environment to support the subsequent scaling up of MMS; (2) ensure delivery platforms that support the introduction of MMS as part of a comprehensive package of maternal nutrition interventions; (3) ensure that pregnant women and healthcare workers understand the importance of MMS and support its use; and (4) integrate MMS monitoring into routine nutrition information systems and generate evidence to inform national scale-up.^9^ Evidence to inform national scale-up will come in part from the two-arm facility-based cluster-randomised trial described here.

### Study design

To address the primary objective of effect on birth weight, continuous data collection of birth weights from all births in enrolled health facilities is carried out between January 2023 – December 2024, also recording data on MMS and IFA exposure amongst the mothers of newborns. To address secondary objectives on processes and costs, this continuous data is supplemented by baseline, midline, and endline sample surveys of ANC users, health facility staff, facility readiness assessments, and qualitative interviews (figure 1). The evaluation team plays no role in the implementation.

**Figure 1.**
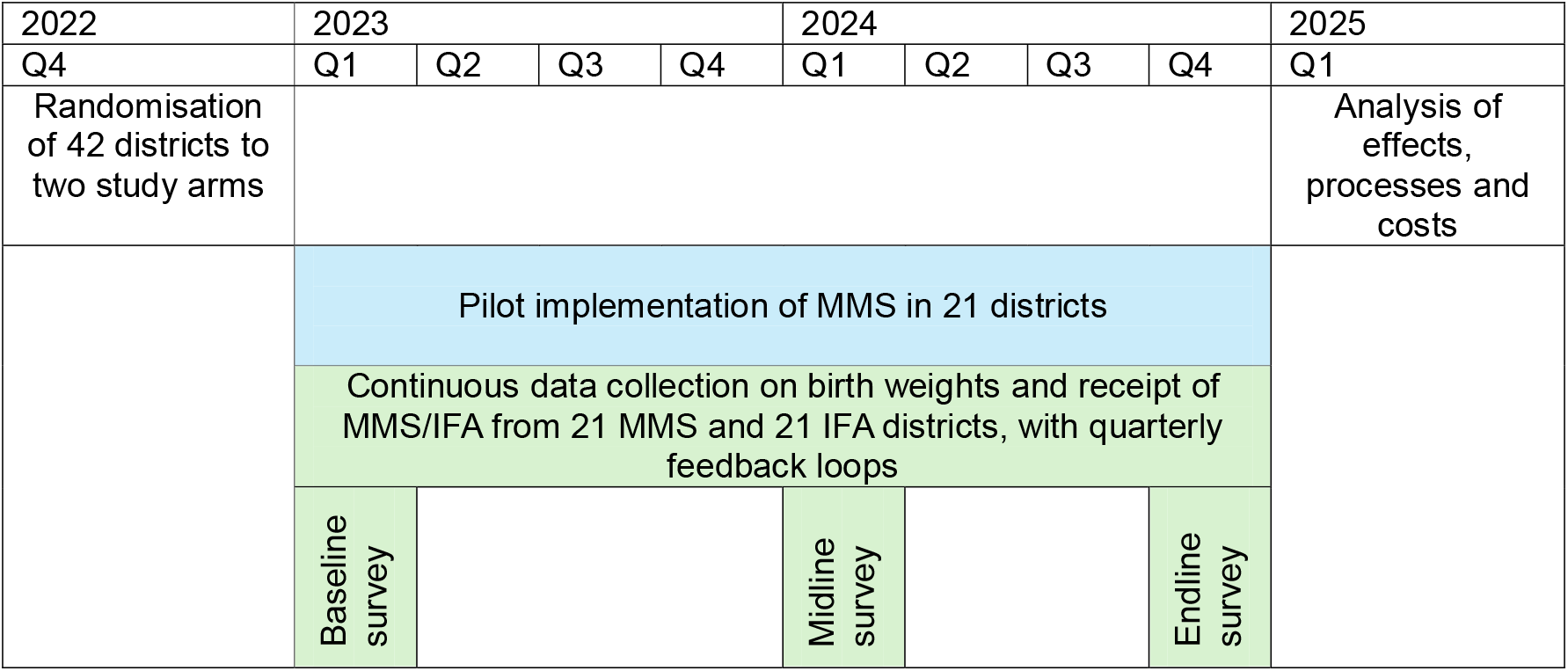
Implementation and evaluation timeline for the MMS pilot in Ethiopia.

### Randomisation

This two-arm trial has 42 districts randomised either to a comparison arm that continues to deliver IFA as part of the standard ANC package, or to an intervention arm where IFA is removed and replaced with MMS (figure 2). Districts were selected as follows. In November 2021 the MoH created a “long list” of 286 districts that were assessed to be secure at that time. From that “long list”, MoH and UNICEF selected a shortlist of 42 districts across five regions where focal persons were present and provided this list to the evaluation team. Stratified randomisation by region was needed to ensure that districts from all five regions be included in both study arms. Districts were therefore listed within regions, assigned a random number then the region-by-region lists ranked by that random number. One half of the districts - those with the lowest ranks - were assigned to the intervention arm and the remaining districts assigned to the comparison arm.

**Figure 2:**
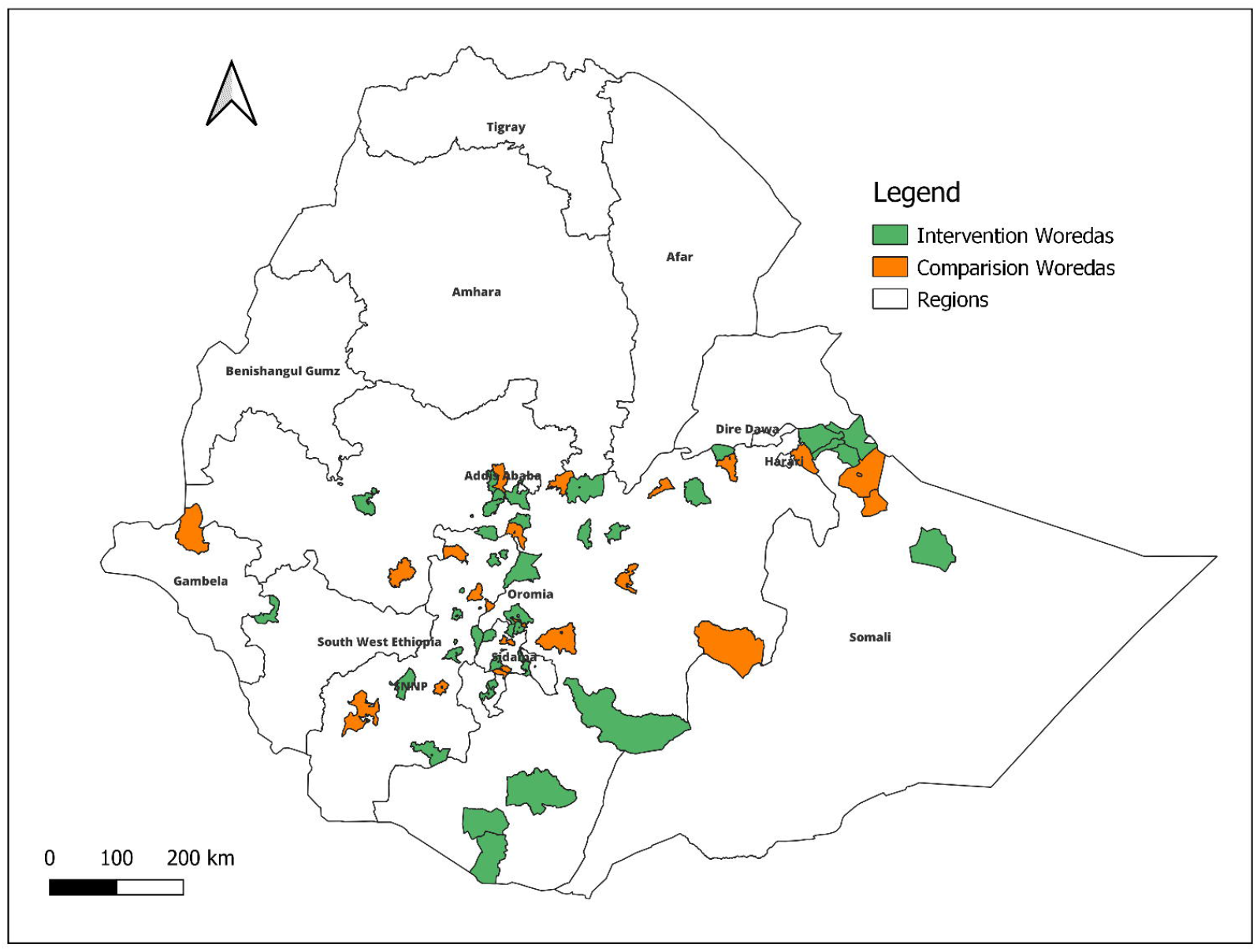
Map of MMS evaluation study area.

### Selection of health facilities

The selection of eligible health facilities within the 42 study districts was conducted as follows. The evaluation team reviewed District Health Information System-2 output to document the volume of births each month in all health facilities across the 42 districts. Within districts facilities were ordered from high to low volume of monthly births and the top five health facilities were visited by the study team for manual record review. Health facilities with at least 15 births per month were enrolled as study health facilities. In total, 44 health facilities were enrolled in the 21-district MMS arm and 54 health facilities were enrolled in the 21-district IFA arm.

### Effect on birth weight: continuous data collection

Continuous data collection of the outcome measure (birth weight) and exposure (receipt and use of interventions) is collected on all eligible births in enrolled health facilities over a period of 2 years from January 2023 to December 2024.

#### Sample size

The sample size calculation for the continuous data collection was based on birth weight. Previous studies suggest a population mean birth weight of approximately 3000g with a standard deviation of 500g, and that the intra-cluster correlation coefficient (ICC) for birth weights within the same woreda ranges between 0.003 and 0.005.^10,11,12^ The Cochrane meta-analysis suggests that babies born to mothers using prenatal MMS have a mean birth weight that is at least 35g higher than babies born to mothers using prenatal IFA.^3^ Assuming the more conservative ICC, a total of 42 clusters i.e. 21 in each group, each contributing 30 observations (birth weights) at each of two-monthly repeat cross-sectional surveys over 24 months (i.e. 15,120 birth weights in total) would provide 80% power to detect a birth weight difference of at least 35g between infants born to mothers offered MMS and those born to mothers offered IFA, assuming a 5% level of significance.^13^

Record review and experience in the study area suggest that 30 births per cluster every two months is feasible in all clusters and that most clusters will exceed this number, giving the study over 80% power.

#### Sample selection for birth weight

The newborn birth weights for all women who give birth in study health facilities during the continuous data collection period are potentially eligible and will be documented, including the following potential reasons for exclusion:

- women who do not give consent for birth weight data to be recorded;
- women who experience a stillbirth;
- women who experience an emergency health event, including risk to their own life or the life of their newborn.

Data on multiple births are collected, although the primary analysis is restricted to singleton live births.

#### Informed consent

Written informed consent is sought from each individual woman before any data is recorded for the purpose of the evaluation (Annex 1). Birth weight is recorded by midwives as part of usual practice, but no data extraction or data collection of any kind occurs until after the mother and infant are settled and away from the delivery room. At that time, trained midwives provide the mother with information about the study purpose, risks and benefits and answer any questions. Women are assured that they have the right to refuse to participate with no adverse consequence to the care provided.

#### Data collection and data entry processes

Amongst women who agree to participate, data on birth weight and mother’s use of antenatal care and exposure to MMS or IFA is collected by the midwives attending births. These midwives are trained and continuously supervised by study supervisors weekly Telegram, WhatsApp chats and Zoom and bi-monthly site visits. Data are recorded on paper forms that have pre-assigned unique numeric identifiers then collected by supervisors for double data entry and verification and stored in a secure room.

#### Variables captured through continuous data collection

A one-page form was co-designed with midwives to capture outcome and exposure data as efficiently as possible, prioritising alignment with existing information recorded by the midwives on women’s health cards. The indicators included on this one-page form are shown in Table 2.

**Table 2:** Indicators available from the continuous data collection one-page form on all births.

| Indicator | Numerator | Denominator |
| --- | --- | --- |
| Maternal characteristics including age, parity, birth outcome, mode of delivery, number and timing of ANC visits. | # women with the characteristic | All women registered who gave consent to participate |
| Newborn characteristics including gestational age at birth, singleton or multiple birth, sex | # newborns with the characteristic | All newborns contributing birth weight data |
| Mean birth weight | All recorded singleton birth weights | N/A |
| Receipt of supplements this pregnancy, disaggregated by IFA/MMS | All women who report having received either MMS or IFA while pregnant | All women whose newborn contributes birth weight data |
| Use of supplements this pregnancy, disaggregated by IFA/MMS | All women who report having consumed any number of tablets while pregnant | All women whose newborn contributes birth weight data |
| Use of at least 90 supplements this pregnancy, disaggregated by IFA/MMS | All women who report having consumed at least 90 tablets while pregnant | All women whose newborn contributes birth weight data and who were first given supplements before the third trimester of pregnancy |

#### Data management and quality control

Each health facility is provided with a digital scale (Seca 384) and batteries. At the start of the study, all staff in the maternity ward plus the facility manager were trained to use the scales, including use of calibration weights. A written standard operating procedure is left at the facility and at each subsequent supervision visit the correct use of digital scales and calibration weights is revised through practical exercises. Also during supervision visits, supervisors review the one-page forms to check completeness and to cross-reference the number of forms against the number of recently delivered women recorded in the facility register: any discrepancies are discussed in-person with facility staff with the aim of continuously improving data quality. On return to Addis Ababa, the one-page forms are submitted for double-data entry and quality control checks are run by data managers for completeness, outliers, and to run histograms to check distribution of birth weight data.

During each facility visit, supervisors aim to make a re-interview home visit to one recently delivered woman who was discharged during the week of the supervision visit and who lives in the neighbourhood of the health facility. If the mother is identified and agrees to be re-interviewed, the supervisor works with the mother to complete the one-page form again then cross-references responses with the original form completed by the midwife. Again, any discrepancies are used to provide feedback to midwives as part of efforts to continuously improve data capture processes. Re-interview one-page forms are also returned for double-data entry and subsequent analysis as matched pairs with the original form completed by midwives.

All data associated with this MMS evaluation is anonymized and stored on secure servers at EPHI. A data sharing agreement exists between EPHI and LSHTM to give equal access to all anonymized datasets and describes dissemination plans to key audiences in Ethiopia and through publication.

#### Statistical analysis for birth weight outcomes

Statistical analysis will be conducted at the individual mother-child dyad level, with appropriate adjustment for clustering within districts. Data on birth weights will be summarised at repeat two-monthly intervals, creating 12 cross-sectional summaries of mean birth weight in each study arm.

For the primary *intention-to-treat* analysis, the mean birth weight of participants will be summarised according to their treatment allocation regardless of their uptake of the intervention. We will use a mixed effects generalised linear model assuming a Gaussian distribution of the outcome with cluster-level random effects to estimate the overall mean difference in birth weight over the whole duration of the evaluation, comparing MMS versus standard care (IFA) clusters using 95% confidence intervals.

For the secondary *per-protocol* analysis, the analysis of mean birth weight will be repeated but in datasets restricted to women (i) who received the interventions within the first 24 weeks of pregnancy and (ii) who self-report consuming at least 90 supplement tablets during pregnancy.

### Process evaluation: facility surveys

A process evaluation^14^ is nested within the overall trial design, focusing predominantly on MMS implementation over time but also drawing comparison with processes for IFA where relevant. Facility surveys are carried out at three points in time (January-April 2023, April-June 2024 and January-February 2025) and are described here. The facility survey involves (i) a health facility readiness assessment, which assesses the availability of essential supplies and services for antenatal care, (ii) interviews with the health staff providing services on the day of survey, to assess their knowledge of the importance of nutritional supplements, (iii) exit interviews with service users, to collect information on receipt and use of nutritional supplements, and (iv) a sub-sample of qualitative interviews with both staff and service users, aimed at exploring attitudes towards nutritional supplements in individuals and their communities. Voluntary written informed consent is sought from all participants.

#### Sample size

Facility surveys are conducted in the same health facilities (health centres and hospitals) included in the continuous data collection. One staff member providing ANC on the day of survey is interviewed per facility. In addition, for each health centre one health post is randomly selected for the facility readiness assessment and health staff interview.

The sample size estimation for exit interviews with facility users is based on detecting a 15 percentage point or greater difference between two point estimates in the acceptability of use of nutritional supplements, assuming a 50% baseline coverage. A sample size of 375 service users per time point, per study arm, will have 80% power to detect a change of 15 percentage points using a 5% significance level, a two-sided test, a design effect of 2.00 and ten percent losses.^13^

The sample sizes for the qualitative interviews are expected to be approximately 20 in each study arm, to be finalised with reference to data saturation after daily summaries.

#### Data management and quality control

Primary data collection tools are designed with input from key stakeholders in English then translated to local languages (Amharic and Oromifa) with back translation to English for consistency checks. Quantitative tools are programmed in Open Data Kit (ODK) using limits on pre-coded responses to minimise errors as well as internal checks that require data input before advancing to another question. All tools are pre-tested before finalisation and used alongside a detailed survey instruction manual during data collection by a survey team who have received classroom training. During data collection, supervisors review data daily before uploading to the secure server from where the data manager also reviews and runs consistency checks on key indicators.

The qualitative data are collected by recording interviews then transcribing and translating. In addition, field notes are made each day to make sure that insights are immediately used to update the interview guides. All data is collected using password protected electronic devices and stored on secured servers.

#### Data analysis

The process evaluation will be descriptive, including questions about the fidelity of MMS implementation (the extent to which the MMS pilot was implemented as planned), the dose of implementation (the availability of MMS and of trained staff in health facilities), adaptation (the extent to which the planned protocol for integration of MMS within routine antenatal care needed to be amended as implementation proceeded, for example because of supply chain or acceptability issues), reach (the extent to which the population in need of MMS are able to access it, and any systematic inequalities therein) and context (the health system structures within which implementation occurs). Regarding women’s knowledge, acceptability and adherence of MMS and IFA, point estimates and their 95% confidence intervals will be calculated. Mean or median days adherence to supplements will be stratified by timing of first ANC attendance. Evidence of change between area or between time point will be determined through chi^2^ tests or t-tests, as appropriate.

For qualitative data, thematic analysis will be carried out. Transcripts will be read for familiarization of conceptual themes to emerge around preferences, enables and barriers to the use of supplements. A coding template and code book will be developed using NVivo and all interviews coded prior to analysis.

### Cost and cost-effectiveness

The study aims to cost MMS and evaluate the incremental cost-effectiveness of MMS compared to IFA as part of routine ANC, based on local costs and birth weight outcomes. Ingredients-based costing will be applied and a decision analytical model employed to estimate the incremental cost-effectiveness of MMS compared to IFA from the health provider perspective (excluding the costs borne by the clients accessing the services). For costs, data collection will be nested in a sub-sample of health facilities included in the midline facility survey and include interviews with health facility managers, district and regional health office managers, MoH staff and implementing partners. If the intervention demonstrates evidence of effectiveness on mean birthweight a full economic evaluation will be carried out to calculate the relative value for money of MMS compared to the IFA standard of care, with methods aligned with the the iDSI Reference Case for Economic Evaluation criteria^15^ and with the Consolidated Health Economic Evaluation Reporting Standards (CHEERS),^16^ and compared with estimates generated using the published MMS Cost-Benefit Tool which draws from global relative effectiveness data.^17^

### Ethics and dissemination

Ethical approval was obtained from the institutional review board of the Ethiopian Public Health Institute [EPHI-IRB-455-2022] and the London School of Hygiene and Tropical Medicine [LSHTM ref 28021]. All participants will be free to participate or not to participate and written informed consent will be sought prior to data collection. Any change to the approved protocol will be communicated in writing to the IRB committees. The results from the study will be disseminated in-country, at conferences, and in a peer-reviewed journal.

### Oversight and monitoring

An independent Data Safety and Monitoring Board (DSMB) was established before the start of the trial to review, with strict confidence, the trial data approximately yearly for the duration of the trial. No formal interim analysis of the data is planned. Information on the rate of stillbirths in health facilities over time are collected and reported to the DSMB.

A steering group for the Ethiopian MMS pilot as a whole is led by the MoH and convenes monthly to bring together implementation and evaluation teams who share progress updates.

Finally, three types of feedback loop link data collected to implementation. First, supervisors alert MoH in the event that any health facility has a stock out of MMS or IFA. Second, data summaries from the one-page form are tabulated every four months and shared with the implementation team to facilitate continuous monitoring and decision making for program improvement. And finally, facility survey reports are shared with the steering group and include requested tabulations that may inform decision making.

## Discussion

This pragmatic trial aims to deliver actionable evidence on the programme effectiveness of a pilot to introduce MMS as the nutritional supplement of choice in Ethiopia’s routine antenatal care package, replacing IFA. Responding to requests from MoH and consistent with WHO guidelines, evidence will be provided on the effect on mean birth weight of changing from IFA to MMS. Additionally the study will assess the costs associated with MMS implementation and evaluate processes that affect successful uptake. Feedback loops are an integral part of the design so that evaluation contributes to the potential for success of this pilot. The trial has three unavoidable but important limitations. First, the need for accurate measures of birth weight requires a facility-based study design, meaning that the study population are likely to be better health-care seekers on average than the general population. Second, the large sample size needed to detect a difference of at least 35 grammes in mean birth weight necessitates working over a very large geographical area, within routine systems, and with health staff rather than posting dedicated study staff in health facilities: continuous close attention is needed on data quality. Finally, the secondary analyses of difference in mean birth weight by exposure to supplements relies on women’s self-reports about tablets received and consumed, and these self-reports are known to have sub-optimal validity.^18^

Nonetheless, this comprehensive study of the effect of an MMS policy shift on health benefits, processes and costs responds directly to country need and will make an important contribution to global learning.

## Supporting information

Supplement_SPIRIT checklist

## Data Availability

This is a protocol manuscript. All data produced in the work described here will be available upon reasonable request to the authors.

## Acknowledgements

We are grateful to the participants of this study for freely giving their time during interview and to the health facility staff and district and regional health management teams who facilitated data collection processes.

## Author statement/ Contributors

This study represents a collaboration between the London School of Hygiene & Tropical Medicine and the Ethiopian Public Health Institute. TM and MT are PI and co-PI and wrote the first draft of the protocol; AD and AKD are research leads; SA and EA lead the costing study; CO, BT and KZ are responsible for data management and trial statistics, LAP and JS are senior researchers on the team; and GT and AT contribute to the process evaluation. All authors read and approved the final version of the manuscript.

## Trial sponsor

London School of Hygiene & Tropical Medicine. Contact

## Patient and Public Involvement Statement

Patients or the public were not involved in the design, or conduct, or reporting, or dissemination plans of our research

